# A chest-CT and clinical chemistry based flowchart for rapid COVID-19 triage at emergency departments – a multicenter study

**DOI:** 10.1101/2020.10.29.20218743

**Authors:** Steef Kurstjens, Eva-Leonne Göttgens, Bob Smit, Bent Postma, Carl Kluge, Armando van der Horst, Eva H.J. Lamboo, Caroline M.M. Janssen – Te Slaa, Robert Herpers, Yvette C.M. Kluiters-de Hingh, Arthur du Mée, Marjan Veuger, Rob van Marum, Peter de Jager, Marc G. L. M. Elisen, Ron Kusters, Martin Schuijt, Marc H. M. Thelen

## Abstract

**Background:** Due to the large number of patients with coronavirus disease 19 (COVID-19), rapid diagnosis at the emergency department (ED) is of critical importance. In this study we have developed a flowchart based on two well-known diagnostic methods: the ‘corona-score’ and the ‘CO-RADS’. This flowchart can be used in hospitals that use chest-CT, instead of chest X-ray, for COVID-19 suspected patients at the ED.

**Methods:** ED patients (n=1904) from the Jeroen Bosch Hospital, Amphia Hospital, HagaHospital, Elisabeth TweeSteden Hospital, Bernhoven Hospital and Slingeland Hospital were included. A laboratory-based ‘corona-score’, without radiology, called the ‘lab-corona-score’ was combined with a chest-CT based radiology scoring system (CO-RADS), to develop a flowchart. The performance was assessed by sensitivity/specificity analyses using the RT-PCR outcome or the physician’s final diagnosis as golden standard.

**Results:** Out of the 1904 patients, 611 (32.1%) patients tested positive for the SARS-CoV-2 virus. The lab-corona-score alone had an AUC of 0.86, a sensitivity of 87% and a specificity of 88% using cut-off values of 0-2 (negative) and 8-10 (positive). Of 255 patients, from the Amphia and Slingeland Hospitals, a CO-RADS score was determined. The flowchart, which combined the ‘CO-RADS’ with the ‘lab-corona-score’, was developed based on data from Slingeland Hospital (sensitivity 97%, specificity 96%). Hereafter, the performance of the flowchart was validated using an independent dataset from Amphia hospital, and reached a sensitivity of 98% and specificity of 93%. A decision could be made in 79% of the patients, which was correct in 95% of the cases.

**Conclusion:** This flowchart, based on radiology (CO-RADS) and clinical chemistry parameters (lab-corona-score), results in a rapid and accurate diagnosis of COVID-19 at the ED.

## INTRODUCTION

The outbreak of the novel SARS-CoV-2 virus in March has resulted in a huge increase in the number of patients at the emergency department (ED). ‘Corona-virus disease 2019’ (COVID-19) is diagnosed by the detection of viral genetic material by real-time reserve transcriptase polymerase chain reaction (RT-PCR). However, it takes considerable time before the result of the RT-PCR is available, in which time the clinicians need to take decisions regarding treatment and isolation protocols. In order to make a well-balanced decision whether a patient is infected with COVID-19, the treating ED physician has to combine the clinical presentation, radiology and clinical chemistry parameters, which therefore require a standardized interpretation

In March 2020 the CO-RADS (COVID-19 Reporting and Data System) was introduced: a method for standardized documentation and risk stratification of a chest-CT for all patients with a suspected COVID-19 infection.^(1)^ The goal of the classification is to improve the communication between physicians, facilitate the comparison of results between institutions and to provide a basis for scientific research. The CO-RADS methodology should be mainly used in a medium-to-high prevalence of COVID-19.^(1)^ In addition, authors of CORADS diagnostics studies have suggested to add routine laboratory parameters to improve the performance of the CORADS classification system.^(2)^

COVID-19 patients often present with a moderately elevated c-reactive protein (CRP), an elevated lactate dehydrogenase (LD), elevated ferritin, lymfocytopenia and a normal neutrophil count. ^(3, 4)^

The interpretation of clinical chemistry parameters is an excellent way to differentiate between a viral and bacterial pneumonia. Based on the situation in March 2020, the ‘corona-score’ was developed (www.corona-score.com). The corona-score has received widespread international attention, and has been validated and implemented in multiple hospitals worldwide.^(5-7)^ The corona-score is a diagnostic algorithm that, based on clinical chemistry parameters, age, sex and a crude interpretation of the chest X-ray, can determine with high accuracy whether a patient at the ED is positive or negative for COVID-19.^(8)^ The scoring system and chosen cut-off points for each variable have been optimized using artificial intelligence. In order to combine the corona-score with the CO-RADS classification, the points for the radiological interpretation have been removed from the corona-score, resulting in the so-called ‘lab-corona-score’.

In this retrospective multicenter research we investigated whether combining the ‘CO-RADS’ with the ‘lab-corona-score’ is an efficient method to distinguish COVID-19 positive from negative patients at the ED.

## METHODS

### Patient population

In this retrospective multicenter study the performance of the ‘lab-corona-score’ separately, and in combination with the CO-RADS score, was investigated. For the performance of the lab-corona-score separately all ED patients were included that received a SARS-CoV-2 RT-PCR test. Patients were excluded if there were missing clinical chemistry parameters due to a (pre-)analytical error, an invalid RT-PCR result or a hemolytic blood sample (Figure 1A). In total 1904 patients were included: Jeroen Bosch Hospital (n=187, 7-28 April 2020), Amphia Hospital (n=377, 1-30 April, 2020), Bernhoven Hospital (n=579, 24 March - 20 April, 2020), Slingeland Hospital (n=104, 5-29 April), HagaHospital (n=429, 23 April - 20 July, 2020) and Elisabeth-TweeSteden Hospital (n=248, 8-30 April, 2020) (Figure 1A).

**Figure 1.**
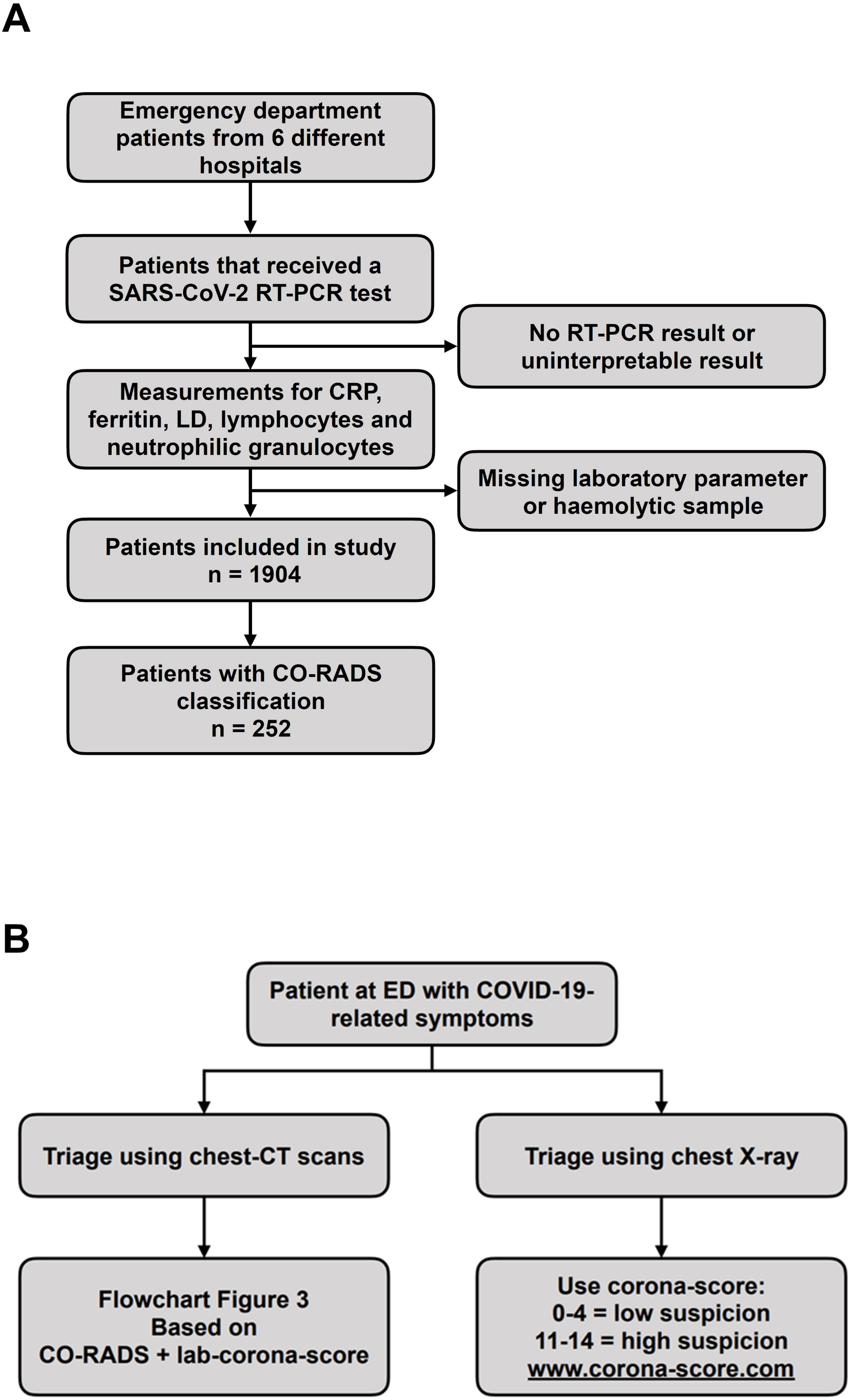
Flow chart of the study setup. (A) Patients from the emergency department (ED) of the Jeroen Bosch Hospital, Amphia Hospital, Slingeland Hospital, HagaHospital, Elisabeth TweeSteden Hospital and Bernhoven Hospital that received an SARS-CoV-2 RT-PCR test, were included. Patients with a missing or non-interpretable RT-PCR result, and patients with missing clinical chemistry parameters, were excluded. In total 1904 patients were included, of whom 252 (from Slingeland Hospital and Amphia Hospital) also had a CO-RADS score. (B) If a hospital uses chest-CT scans for the triage at the ED, the left arm of the flow chart can be used. If the ED uses chest X-ray, the corona-score can be used.

### Laboratory measurements

Venous blood was drawn from all patients that presented at the ED with symptoms that could be related to COVID-19. A nasopharyngeal or oropharyngeal swab was taken for the SARS-CoV-2 RT-PCR test. These were performed by the department of medical microbiology at the institute using the following methods: Thermofisher 7500SDS Thermofisher and Roche cobas-6800 for E-and RdRP gene (Amphia Hospital), Cepheid Xpert Xpress SARS-CoV-2 on a GeneXpert DX System (Slingeland Hospital and emergency tests from HagaHospital), In-house kit on an Applied Biosystems 7500 RT-PCR System (HagaHospital), PRD-06419 Aptima SARS-CoV-2 on a LIAISON Panther System (Jeroen Bosch Hospital and Bernhoven Hospital).

Clinical chemistry and hemocytometric tests were measured in the venous sample from the initial presentation at the ED using the following analysers: Siemens Advia Chemistry XPT and Advia 2120i (Jeroen Bosch Hospital), Roche cobas-8000 and Sysmex XN-6000 (Amphia Hospital), Dimension Vista 1500 and CELL-DYN Sapphire (Bernhoven Hospital), Roche and Sysmex (Elisabeth TweeSteden Hospital), Roche cobas-6000 and Sysmex XN-1000 (HagaHospital), and Roche CobasPro and Sysmex XE-5000 (Slingeland Hospital). Due to the large variation in the ferritin measurements between the platforms, the ferritin concentration by Roche was used as a reference. Ferritin measurements were harmonized by multiplying the concentrations measured by Siemens analysers by a factor of 1.2 ^(8)^.

### Lab-corona-score

In order to combine the ‘corona-score’ with the CO-RADS classification we removed the interpretation of the chest X-ray, resulting in the so-called ‘lab-corona-score’. In this score, points are attributed to each parameter as described in Table 1. The ‘lab-corona-score’ is trimmed from a minimum score of 0 to a maximum score of 10.^(1)^ The result of this ‘lab-corona-score’ can be easily implemented into the electronic laboratory system, so that it is automatically reported directly into the patient file. More information can be found at www.corona-score.com.

**Table 1:** Point-based system for the lab-corona-score. The total score is cut-off, with 0 as a minimum score and 10 as a maximum score.

| Variable | Points |  |  |  |  |  |  |
| --- | --- | --- | --- | --- | --- | --- | --- |
| **Age (years)** | ≤75 | 76-79 | ≥ 80 |  |  |  |  |
| Points | 0 | 1 | 2 |  |  |  |  |
| **Sex** | Female | Male |  |  |  |  |  |
| Points | 0 | 1 |  |  |  |  |  |
| **CRP (mg/L)** | 0–9 | 10–14 | 15–38 | 39–69 | 70–193 | 194–303 | ≥304 |
| Points | 0 | 1 | 2 | 3 | 2 | 1 | 0 |
| **Ferritin (µg/L)** | ≤15 | 16–179 | 180–301 | 302–538 | ≥539 |  |  |
| Points | -1 | 0 | 1 | 2 | 3 |  |  |
| **LD\* (U/L)** | ≤257 | 258–265 | 266–397 | ≥398 |  |  |  |
| Points | 0 | 1 | 2 | 3 |  |  |  |
| **Lymphocytes (10<sup>9</sup>/L)** | ≤1.2 | ≥1.3 |  |  |  |  |  |
| Points | 1 | 0 |  |  |  |  |  |
| **Neutrophilic granulocytes (10<sup>9</sup>/L)** | ≤5.1 | 5.2–7.9 | 8.0–9.0 | 9.1–10.3 | ≥10.4 |  |  |
| Points | 0 | -1 | -2 | -3 | -4 |  |  |

CRP, c-reactive protein; LD, lactate dehydrogenase, \*the result was multiplied by a factor of 0.9 in case of a mildly hemolytic sample.

### Radiology

The chest-CT images were interpreted and scored according to the CO-RADS classification by the radiologists of the Amphia and Slingeland Hospitals.^(1)^ The CO-RADS classification is scored from 1 to 6 (Table 2). A CO-RADS score of 1 is the lowest grade of suspicion, fitting a normal chest CT-image or a non-infectious image. A CO-RADS score of 5 is the highest grade of suspicion, in which chest CT-findings include typical signs of COVID-19, such as multifocal and bilateral ground glass consolidations which extend into the pleura, with or without consolidations. ^(1, 9)^ Low dose CT-scans were performed using a Siemens Somatom AS (Amphia Hospital) and a Siemens Somatom Flash CT (Slingeland Hospital).

**Table 2.** Description of CO-RADS classification

|  | Odds of having COVID-19 | CT-outcomes |
| --- | --- | --- |
| CO-RADS 0 | Unable to assess | For example: low quality images due to artefacts |
| CO-RADS 1 | Very low | Normal CT-images, or evidently non-infectious characteristics. |
| CO-RADS 2 | Low | Images infectious characteristics, however not typical for COVID-19. |
| CO-RADS 3 | Moderate/Dubious | Abnormalities fitting with a viral infection, unclear whether COVID-19 related. |
| CO-RADS 4 | High | Abnormalities fitting with a COVID-19 viral infection, with a possibility of a different etiology. |
| CO-RADS 5 | Very high | Highly typical infectious COVID-19 characteristics. |
| CO-RADS 6 | RT-PCR SARS-CoV-2 positive | Any image |

### Development of the flowchart

For the development of the flowchart (Figure 3), based on the lab-corona-score in combination with the CO-RADS score, data from hospitals were used that perform routine CT-scans at the ED for each COVID-19 suspected patient. Data from Slingeland Hospital (n=104) was used to design the flowchart. In designing this flowchart, we evaluated for which CO-RADS scores the ‘lab-corona-score’ could provide a contribution in making the correct diagnosis. The final decision of the treating physician was used as a golden standard. In 5 patients with a negative RT-PCR result, the treating physician concluded that the RT-PCR result was a false-negative, and the patient was registered as COVID-19-positive. To validate the flowchart an independent dataset was used from the Amphia Hospital (n=148). The result of the RT-PCR was used as golden standard.

**Figure 2.**
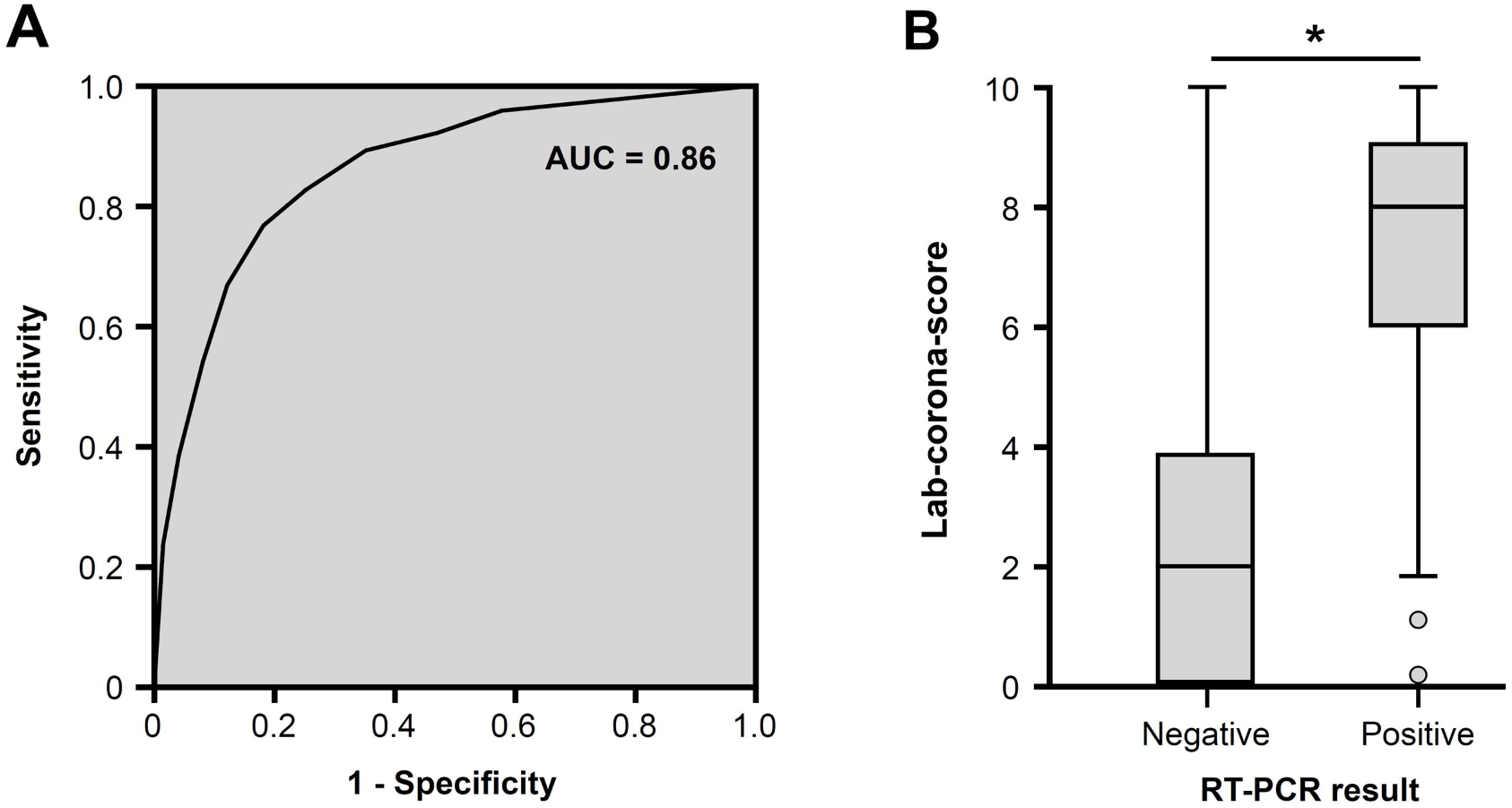
Diagnostic characteristics of the lab-corona-score. In the ‘lab-corona-score’ the points for the interpretation of the radiological image have been removed from the corona-score. (A) The ROC curve of the ‘lab-corona-score’, in which the area under the curve (AUC) is 0.86. (B) Box-plots of the ‘lab-corona-score’ in patients with a negative and positive SARS-CoV-2 RT-PCR result.

**Figure 3.**
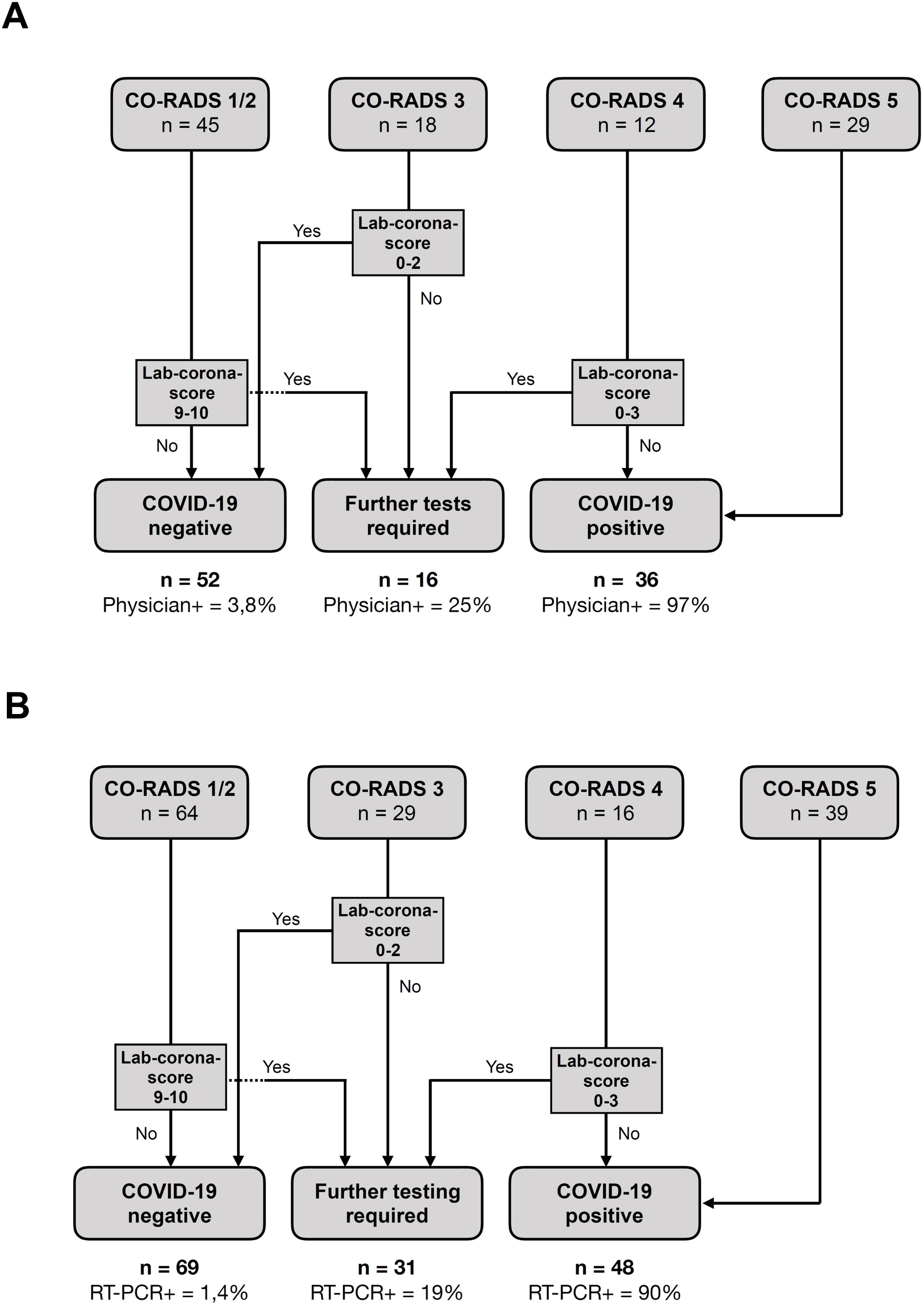
Diagnostic flow chart for COVID-19-suspected patients at the emergency department. (A) The flowchart was developed based on 104 patients of the Slingeland Hospital. The final conclusion of the treating physician was used as golden standard to determine whether a patient was considered COVID-19 negative or positive. (B) Hereafter, the flowchart was validated using an independent dataset of 148 COVID-19-suspected patients of the Amphia Hospital. The result of the RT-PCR was used as golden standard to determine whether a patient was considered COVID-19 negative or positive.

### Statistics

Data were analyzed using Excel 2010 (Microsoft Corporation, VS) and SPSS (IBM, Version 25.0, VS). Lab-corona-scores were compared between the two groups using an unpaired t-test with a Welch correction. A *p*-value of < 0.05 was considered statistically significant.

### Ethical statement

This study was conducted according to the declaration of Helsinki and Guidelines for Good Clinical Practice. The execution of this retrospective observational study of patient records was approved by the regional Medical Research Ethics Committee (METC Brabant), which declared that the use of anonymized patient data is not subject to the regulations of the WMO (Dutch Medical Research Involving Human Subjects Act). Consent was obtained by an opt-out agreement.

## RESULTS

In order to combine the ‘corona-score’ with the CO-RADS classification, the radiological interpretation has been removed from the corona-score, resulting in the so-called ‘lab-corona-score’ (Table 1). The original corona-score can be used if chest X-rays are routinely used at the ED instead of chest CT-scans (Figure 1B). The performance of the ‘lab-corona-score’ has been retrospectively analyzed based on 1904 patients from 6 different hospitals. The prevalence of COVID-19 positive patients at the ED ranged from 10% (HagaHospital) to 59% (Bernhoven Hospital, Table 3). Using cut-off values for the ‘lab-corona-score’ of 0-2 (negative) and 8-10 (positive), the sensitivity was 87% and the specificity 88% (Table 3). In this manner, a decision could be made in 61% of the patients. The area under the receiver operating characteristic (AUROC) curve was 0.86 (Figure 2A). The median of the ‘lab-corona-score’ in the COVID-19 negative patients was 2 (n=1293) and in the COVID-19 positive patients 8 (n=611, Figure 2B, *p*<0,001).

**Table 3.** Diagnostic accuracy of the lab-corona-score per hospital. In the lab-corona-score, only the laboratory parameters contributed to the scoring system. The sensitivity and specificity of the lab-corona-score was determined for the individual hospitals, as well the fraction of patients that could be classified. In this instance, a score of 0-2 was judged as negative, while a score of 8-10 was judged as positive. Other cut-offs could lead to a higher sensitivity or specificity, or could be used to classify a higher number of patients.

| Hospital | # Patients | RT-PCR<br>positive<br>(prevalence) | Sensitivity | Specificity | Received<br>classification |
| --- | --- | --- | --- | --- | --- |
| Amphia Hospital | 363 | 23% | 87% | 89% | 58% |
| Bernhoven Hospital | 579 | 59% | 88% | 84% | 67% |
| Elisabeth-TweeSteden<br>Hospital | 248 | 21% | 83% | 93% | 63% |
| HagaHospital | 423 | 10% | 88% | 90% | 58% |
| Jeroen Bosch Hospital | 187 | 28% | 78% | 79% | 50% |
| Slingeland Hospital | 104 | 39% | 91% | 86% | 62% |
| Total | 1904 | 32% | 87% | 88% | 61% |

The flowchart was developed based on data of Slingeland Hospital (n=104). Patients were classified based on their CO-RADS score (Figure 3A). Patients with a CO-RADS of 1 or 2 were classified as COVID-19 negative, except if their ‘lab-corona-score’ was 9-10. For patients with a CO-RADS score of 3, patients were considered negative if their ‘lab-corona-score’ was 0-2. Patients with a CO-RADS score of 4 were classified as COVID-19 positive, except if their ‘lab-corona-score’ was 0-3. All patients with a CO-RADS score of 5 were considered COVID-19 positive. The flowchart obtained a sensitivity of 97% and a specificity of 96%. The flowchart could make a decision in 85% of the patients, which was correct 97% of the times. In the 16 patients where no decision could be given, 4 patients were COVID-19 positive. These all had a CO-RADS score of 3 with a ‘lab-corona-score’ between 4 and 10.

The performance of this flowchart was validated on an independent dataset of Amphia Hospital (n=148, Figure 3B). In this analysis the result of the RT-PCR was used as golden standard. If a patient that initially tested negative for COVID-19 tested positive within 7 days after their initial visit, this patient was considered COVID-19 positive. Nonetheless, it is possible that this population still contains a number of false-negative results due to incorrectly performed swabs. When running this validation-population through the flowchart, 69 (47%) patients were considered COVID-19 negative, of which 1 patient (1.4%) had a positive RT-PCR test. In the flowchart 48 (32%) patients were considered COVID-19 positive, of which 90% had a positive RT-PCR. In conclusion, the validation of the flowchart reached a sensitivity of 98% and a specificity of 93%. In 79% of the patients a decision could be made, which corresponded with the RT-PCR result in 95% of the patients. In the 31 patients where no conclusive decision could be reached by the flowchart, 6 patients tested positive for SARS-CoV-2. Of these 6 patients, 5 patients had a CO-RADS of 3 (lab-corona-score between 3 and 7) and 1 patient had a CO-RADS of 4 (lab-corona-score of 0). The ‘lab-corona-score’ is significantly different between COVID-19 positive and negative patients with a CO-RADS score of 3 or 4 (*p*<0.05, Figure 4A-B), demonstrating the additive diagnostic value of the ‘lab-corona-score’ in combination with radiology.

**Figure 4.**
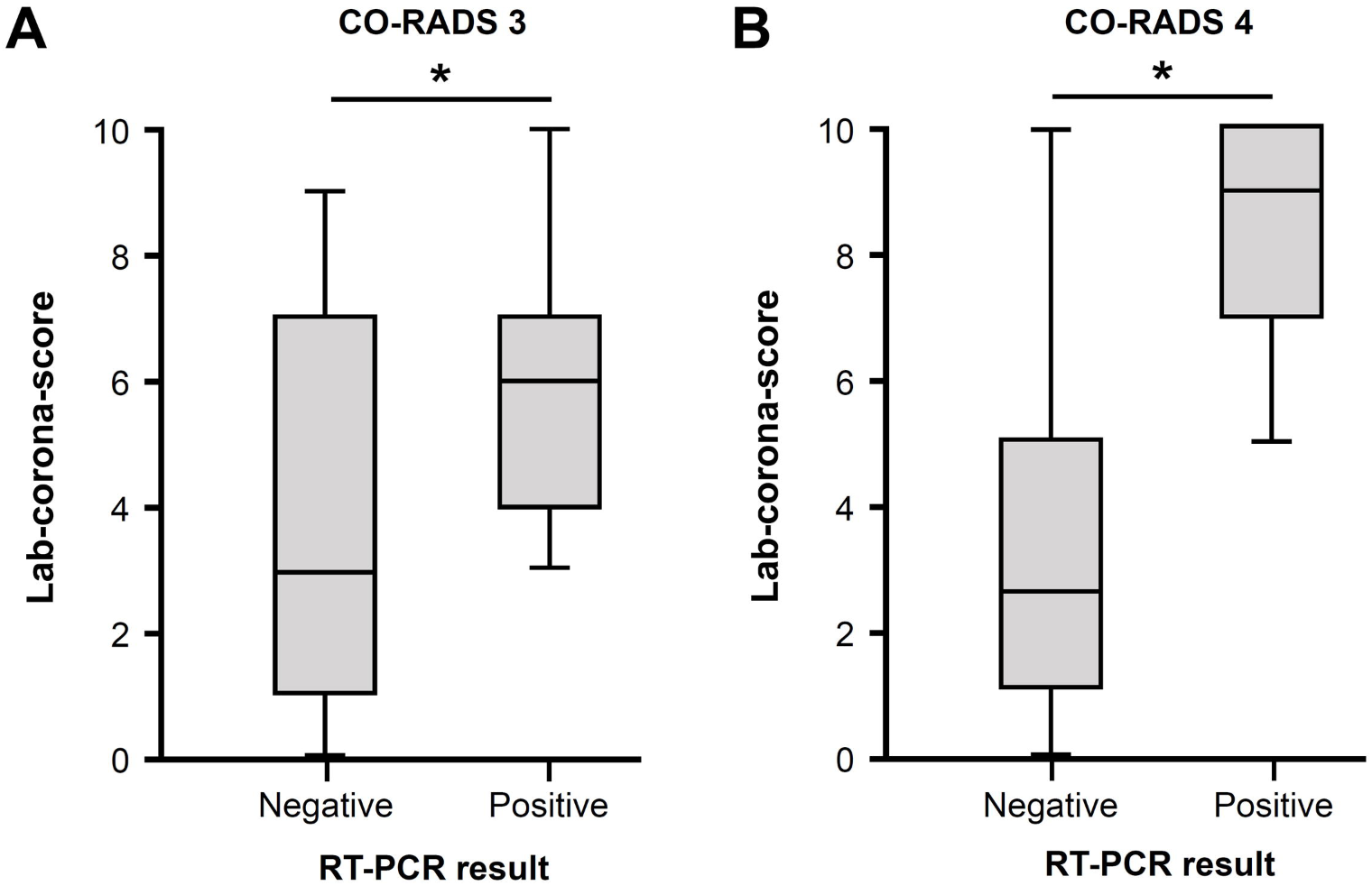
The added value of the lab-corona-score in CO-RADS 3 and 4. Boxplots of the ‘lab-corona-score’ in patients from the Slingeland Hospital and Amphia Hospital who had a CO-RADS of 3 (A) or 4 (B). The ‘lab-corona-score’ is significantly different between the COVID-19 negative and positive patients with a CO-RADS of 3 or 4 (*p*<0.05).

## DISCUSSION

The total diagnostic accuracy of the flowchart is 95-97% and is able to diagnose approximately 80% of patients, using chest-CT and routine laboratory parameters. The proposed flowchart allows for a fast and accurate indication whether patients at the ED with COVID-19 related symptoms are infected with the SARS-CoV-2 virus or not.

Multiple flowcharts have been proposed to support and improve triaging of COVID-19 suspect ED patients. For example, the Dutch Association of Medical Specialists employs a flowchart based on CO-RADS classification, but the flowchart does not take advantage of changes in clinical chemistry parameters in COVID-19 infected patients. In addition, the use of flowcharts that rely on the outcome of SARS-CoV-2 RT-PCR tests are both time-consuming and complicated by the occurrence of false-negative RT-PCR outcomes. Another research group has proposed the use of LD and lymphocyte count as a means to assess the probability of a SARS-CoV-2 infection.^(10)^ However, the use of merely these two parameters raises the possibility of misclassification. For example, LD will always be relatively high in a patient suffering from hemolysis and lymphopenia has numerous pathophysiological causes as well. By combining a larger set of multiple laboratory parameters, this effect will be largely diluted. From our results it is clear that using the lab-corona-score and the CO-RADS classification provides added value as compared to using CO-RADS alone. In this instance a high lab-corona-score will confirm the diagnosis if CO-RADS is 4, but will it often rightfully cast doubts in case of a low lab-corona.

Our study has several strengths. To start with, the study contained data from six independent Dutch hospitals that all calculated and/or reported the corona-score or lab-corona-score. The development and validation of the flowchart was performed on two independent datasets from different hospitals. This strongly contributes to the validity and robustness of the diagnostic performance of the flow chart. Secondly, the lab-corona-score is easy to implement in the laboratory information system. It can be readily reported to the hospital information system, where it will be accessible to all emergency physicians. This, however, will require that a standardised set of laboratory parameters will be requested by ED professionals in order to calculate the (lab-)corona-score. Lastly, the use of this flowchart is significantly faster than current RT-PCR tests, and can therefore reduce both triage time and time to start treatment.

This study also has a set of limitations. During the development of the flowchart, the medical diagnosis of the physician was used as the golden standard. However, in the validation of the flowchart, the results of the RT-PCR were used as golden standard. Because false-negative RT-PCR results as a result of sampling are not uncommon, it is possible that a number of COVID-19 patients have been incorrectly classified as negative. This could have led to an underestimation of the diagnostic performance of the flow chart. A second limitation is that the use of the flowchart requires the performance of a CT-scan for all suspected patients. If a hospital is unable to perform a CT-scan, they will not be able to use this flowchart. In that case, we would like to recommend the use of the original corona-score, with cut-off values of 0-4 and 11-14.^(8)^ A third limitation might be that the diagnostic accuracy of the flowchart might be affected by geographical location and population characteristics, as this study was performed in Dutch hospitals only.^(7)^ Therefore we recommend that hospitals perform a small local validation of the performance of the flowchart prior to implementation. Lastly, as of now we have not been able to whether the performance of the flowchart would be diminished when a large portion of the patients is infected by season flue (influenza). However, it is unlikely that influenza leads to such drastic changes in clinical chemistry parameters, such as ferritin and LD, as seen in COVID-19 infections.

Based on the results of this study we conclude that a flowchart based on a standardised lab-corona-score and radiological CO-RADS classification is an accurate and fast diagnostic tool. Because results of CT-scans and laboratory parameters are rapidly available, it is possible to expeditiously assert the risk of a SARS-CoV-2 infection in EDs. Moreover, the diagnostic tool also reflects the chance of a RT-PCR test result possibly being false-negative. Using this flowchart it is possible to quickly triage COVID-19 suspected patients, and decide on isolation measures.

## Data Availability

Data and further information are available upon reasonable request

## DECLARATION OF INTEREST

Declarations of interest: none

## DATA AVAILABILITY STATEMENT

Data and further information are available upon reasonable request

## FUNDING

None

## Notes

### Competing Interest Statement

The authors have declared no competing interest.

### Funding Statement

No funding

